# Genotype-microbiome-metabolome associations in early childhood, and their link to BMI and childhood obesity

**DOI:** 10.1101/2023.11.13.23298467

**Authors:** Andrea Aparicio, Zheng Sun, Diane R. Gold, Augusto A. Litonjua, Scott T. Weiss, Kathleen Lee-Sarwar, Yang-Yu Liu

## Abstract

The influence of genotype on defining the human gut microbiome has been extensively studied, but definite conclusions have not yet been found. To fill this knowledge gap, we leverage data from children enrolled in the Vitamin D Antenatal Asthma Reduction Trial (VDAART) from 6 months to 8 years old. We focus on a pool of 12 genes previously found to be associated with the gut microbiome in independent studies, establishing a Bonferroni corrected significance level of p-value < 2.29 × 10^−6^. We identified significant associations between SNPs in the FHIT gene (known to be associated with obesity and type 2 diabetes) and obesity-related microbiome features, and the children’s BMI through their childhood. Based on these associations, we defined a set of SNPs of interest and a set of taxa of interest. Taking a multi-omics approach, we integrated plasma metabolome data into our analysis and found simultaneous associations among children’s BMI, the SNPs of interest, and the taxa of interest, involving amino acids, lipids, nucleotides, and xenobiotics. Using our association results, we constructed a quadripartite graph where each disjoint node set represents SNPs in the FHIT gene, microbial taxa, plasma metabolites, or BMI measurements. Network analysis led to the discovery of patterns that identify several genetic variants, microbial taxa and metabolites as new potential markers for obesity, type 2 diabetes, or insulin resistance risk.

## 1. Introduction

Our understanding of human genetics through genomic sequencing has progressed immensely over the past decades and key advances in genome-wide association studies (GWAS) have played an important role in the development and improvement of treatments and medications [1,2]. On the other hand, the human microbiome (i.e., trillions of microbes and their genes that coexist with us) is known to influence human health and diseases [3,4]. In particular, many interactions between the gut microbiome and the immune system, inflammatory processes, and cardiovascular and mental health conditions have been found [5]. Further, alterations to the healthy microbiome, also known as dysbiosis, have been associated with many conditions such as metabolic disease and obesity [6–8].

The gut microbiome has been thought to be acquired during birth [9] and shaped predominantly by the environment throughout the host’s life [10,11]. However, there is growing evidence that points towards the heritability of certain components of the gut microbiome [12] through microbiome-GWAS (mGWAS). Examples of heritable taxa associated with changes in BMI or obesity that these studies have identified are the species *Christensenella minuta* [13], the archaeon *Methanobrevibacter smithii,* the genus *Blautia,* and *Akkermansia muciniphila* in the Verrucomicrobia phylum [14].

Due to the very large number of variants in the human genome, and of microbial species found in the gut microbiome, statistical power is one of the biggest challenges in mGWAS. Although some efforts have been made to revisit (and relax) the significance threshold for GWAS [15–17], the strict p-value < 5 × 10^!8^introduced almost three decades ago [18] is still widely used. Additionally, microbiome datasets are usually highly sparse [19], and a high diversity among individuals has been found [20]. As a result, only a small proportion of species is shared among individuals and, to a greater extent, among populations. This has led to very few mGWAS associations surviving multiple testing correction, and even less replication among different studies. However, genetic variants in a limited set of genes have been consistently found to be significantly associated with microbiome features in multiple independent studies. Examples of these are the associations of the lactase (LCT) gene locus with the genus *Bifidobacterium* found in five independent studies, and microbiome associations of the alpha 1-3-N-acetylgalactosaminyltransferase and alpha 1-3-galactosyltransferase (ABO) gene found in German, Dutch, and Finnish cohorts [21].

Multi-omics studies offer a unique opportunity to unravel the complex mechanisms that govern health and disease from an integral perspective, paving the way towards precision medicine [22]. However, multi-omics data integration presents its unique set of challenges such as a further increase in the data dimensionality, and interpretability of the results [23]. Metabolites are often seen as the output of many biological processes and have been called “the link between genotypes and phenotypes” [24]. Results of metabolomics studies have found associations with conditions such as Alzheimer’s, type 2 diabetes, and obesity, making metabolites good biomarkers for these ailments [25]. Yet, very few multi-omics studies have studied the individuals’ genotype, microbiome, and metabolome simultaneously. Notable examples aimed to understand the combined effect of the microbiome, diet, and genetics on individuals’ metabolome with a wide range of objectives such as designing a diet that promotes overall homeostasis [26], or understanding the mechanistic processes in colorectal cancer [27].

Early life physiology can have a long-term impact on human health. Changes that occur during childhood can predispose individuals to negative outcomes later in life. An accumulating number of studies have shown that factors that disrupt the gut microbiome of infants and young children can have implications for health outcomes such as asthma, allergies, obesity, and gastrointestinal conditions [28]. Similarly, it has been suggested that early life metabolomics could shed light on the mechanisms that lead to childhood obesity [29], autism [30], and IBD [31], among other conditions. The early-life microbiome and metabolome have been studied simultaneously a few times in recent years, often focusing on specific health outcomes such as inflammation [32,33], asthma [34,35], or diet [36,37]; an effort to characterize the microbiome-metabolome relationship in early life found limited inter-omic concordance in children up to 12 months of age [38].

Here, we performed a multi-omics study to reveal associations between the genotype, microbiome, and metabolome of a cohort of 676 children in the longitudinal Vitamin D Antenatal Asthma Reduction Trial (VDAART), a multi-site randomized, double-blind, placebo-controlled trial of vitamin D supplementation during pregnancy. VDAART is an ongoing study and the offspring of the women originally enrolled have been followed for 10 years; blood and fecal samples, and anthropometric measurements have been collected at several time points. To address the challenge of high dimensional data, we took a knowledge-based sequential approach to select the variables to include in our analysis. We first identified a subset of 10 SNPs in the FHIT gene, out of a pool of 12 genes known to be associated with microbiome features, that are significantly associated with microbiome richness and composition and children’s BMI, using a Bonferroni-corrected significance threshold. Then, we identified a set of 6 genera that are differentially abundant with respect to genotype in the set of 10 FHIT SNPs. Finally, we looked for simultaneous associations between the genera and FHIT SNPs of interest, and the children’s metabolome. This approach allowed us to select the variables to include in detailed analysis for every omics data type, therefore reducing the dimensionality of the data and the testing burden. Our main results include two triple and one quadruple associations (where variables of different omics are significantly associated in a coherent directional manner with each other). Notably, we found that a homozygous minor genotype in the rs34723559 locus is associated with the enrichment of the insulin inhibitor sphingomyelin, and increased BMI. This result could help expand the known genetic markers for obesity and obesity-related complications such as insulin resistance and type 2 diabetes.

## 2. Materials and methods

### 2.1. VDAART and Children’s Characteristics

We analyzed data from participants in the VDAART clinical trial [39]. VDAART is a randomized controlled trial of Vitamin D supplementation during pregnancy to prevent asthma in offspring conducted in the United States (St. Louis, Boston, and San Diego; NCT00920621). The study protocol was approved by the institutional review boards at each participating institution and at Brigham and Women’s Hospital. All participants provided written informed consent [40]. The stool and blood samples were collected at three different sites: Boston, MA; San Diego, CA; and St. Louis, MO. The available subjects’ characteristics from the initial enrollment questionnaire are the family household income, mother’s education level, and parents’ and children’s race and ethnicity. Race and ethnicity information was originally collected in VDAART, because they are determinants of the circulating 25-hydroxyvitamin D levels; the race and ethnicity of parents was self-reported and that of every child participant was reported by their parent. Participants were asked to first categorize themselves and their child as either Hispanic or non-Hispanic, then to categorize their race into prespecified categories. Race/ethnicity (called “race” hereafter) groups were collapsed into 3 groups for the analysis: Black or African American (called “Black” hereafter), White, non-Hispanic (called “White” hereafter), and Hispanic and children of other races (called “Other race” hereafter).

### 2.2. BMI measurements

At every yearly visit from age 2 through 8 years, the weight and height of children participating in VDAART was recorded. Child BMI was then calculated as weight in kilograms divided by the square of height in meters. BMI percentiles were calculated using the R package childsds [41]. Children were classified according to their BMI Weight Status Categories (given by the Center for Diseases Control and Prevention [42]), called BMI categories hereafter, as: underweight, with a BMI less than the 5^th^ percentile; normal weight, with a BMI on the 5^th^ percentile or more and less than the 85^th^ percentile; overweight, with a BMI on the 85^th^ percentile or more and less than the 95^th^ percentile; and obese, with a BMI equal or garter than the 95^th^ percentile. Hereafter, we use the term BMI measurements to generically refer to either one of BMI percentiles or categories.

### 2.3. Metabolome profiling

Untargeted metabolomic profiling was performed on plasma collected at ages 1 and 3 years [43]. Metabolomic profiling, mass spectrometer platforms, sample extraction and preparation, instrument settings and conditions, and data handling were performed at Metabolon (Research Triangle Park, NC) [44]. Results were expressed as relative abundance. Based on the assumption that missingness is due to low signal intensity, missing values were replaced with half of the minimum relative abundance observed for the metabolite in question [45]. Finally, relative abundances were log _10_ normalized and Pareto-scaled (mean-centered and divided by the square root of the standard deviation).

### 2.4. Stool samples

At ages 0.5, 1, 3 and 4 years old, (n=256, n=436, n=506, n=314, respectively) child participants of VDAART provided a stool sample. Their parents were asked to collect a 0.5 teaspoon-sized sample 1 to 2 days before a study visit and store the sample in a home freezer before transport with a freezer pack to the study site. Stool was not collected if participants had used antibiotics in the past 7 days. After delivery to the study site, stool samples were immediately stored at -80°C. Microbiome profiling was performed by sequencing the 16S rRNA hypervariable region 4 (V4 515F/816R region) on the Illumina MiSeq platform at Partners Personalized Medicine (Boston, MA).

### 2.5. Genotype principal components and minor allele count

Genotyping was performed in VDAART participants using the Illumina Infinium HumanOmniExpressExome Bead chip. Imputation was performed using Minimac [46] on the Michigan Imputation Server with the 1000 Genomes reference panel [47]. Genotype principal components analysis (PCA) was performed using LASER, which analyzes sequence reads of each sample and places the sample into a reference PCA space constructed using genotypes of a set of reference individuals [48]. Minor allele count of SNPs was obtained using PLINK version 1.9 [49].

### 2.6. Candidate genes

We selected candidate genes for a targeted analysis based on a comprehensive literature review. The criteria for selection employed is that variants in every gene must have been associated with microbiome features in at least two independent previously published studies [21]. This yielded a pool of 12 candidate genes: DMRTB1 in chromosome 1; LCT in chromosome 2; CNTN6, FHIT, and NMNAT3 in chromosome 3; LINC02226 and CTNND2 in chromosome 5; SH3BGRL2 in chromosome 6; TMEM106B in chromosome 7; ABO in chromosome 9; CD5 in chromosome 11; and TMEM132C in chromosome 12. References and details can be found in Table SIxx.

### 2.7. Microbiome composition and principal components

Microbiome samples were centered log-ratio transformed on only their non-zero values, and dimensionality reduction was performed through Robust PCA using the Python package Gemelli, which implements a robust Aitchison PCA [50]. We used the function auto-rpca, which reduced the microbiome composition to three principal components.

### 2.8. Statistical analysis

Unless specifically noted, we used a cut-off p-value of 0.05 to deem results statistically significant for all our analyses. All association analyses were adjusted for *a priori* selected potential covariates: participants’ race, sex, and study site, unless otherwise specified.

#### 2.8.1. Alpha diversity

Alpha diversity measures are estimates of an individual sample’s taxonomic diversity. We computed the observed richness, Shannon, and Simpson indices using the Phyloseq package in R [51]. The observed richness simply counts the number of different taxa present in each sample.

The Shannon and Simpson indices incorporate the measures of richness and evenness of every sample.

#### 2.8.2. Genome-wide association studies

Using PLINK’s association analysis functionality, we applied covariate-adjusted genome-wide association tests to the 21834 variants in the candidate genes, and the three principal components of microbiome composition at every available age, and alpha diversity measures. Accounting for the total number of variants in the candidate genes, a Bonferroni-corrected significance level was established at p-value < 2.29 × 10^−6^, and a suggestive level at p-value < 10^−5^.

#### 2.8.3. Linear regression and mixed models

We used covariate-adjusted linear regression models to search for associations between child genotype and BMI. We fit a linear model for every available BMI measurement (i.e., for BMI each year between ages 2 and 8 years), and deemed an association consistent and significant if it produced a p-value < 0.01 for five or more consecutive BMI timepoints, and the association maintained the same direction for all timepoints across both BMI measurements. We also used covariate-adjusted linear models to search for associations between the relative abundance of children’s metabolites and BMI, genotype, and genera relative abundance. We deemed associations significant if the fit model produced a p-value < 0.01. To search for longitudinal associations between the relative abundance of individual genera and BMI we fit linear mixed models introducing a random intercept for every subject. Linear models were implemented using the lm R function, and linear mixed models were implemented using the function lmer in the R package lme4.

#### 2.8.4. Differential abundance

To search for associations between the subject genotype and BMI, and the abundance of specific taxa, we used MaAsLin2 and its corresponding R package [52], treating the minor allele frequency at a particular locus as a continuous variable. MaAsLin is a multivariate association method that uses additive linear models to detect associations between specific groups and the abundance of taxa, simultaneously treating all the present taxa as outcomes. We used the false discovery rate (FDR) method to adjust the *p*-values for multiple comparisons. The association analyses were performed on genus-level data.

#### 2.8.5. Network integration

Using the results of the association analysis, we built a network using the R package *igraph* where the weight of the edges or links (-1 or 1) represents the direction of the association, and every node or vertex is a variable used in the association studies. Here we use the term loop to refer to a connected component (a set of nodes where any of them can reach any other by traversing edges) with the minimal number of edges. We refer to a loop with congruent edge weights (representing directional associations) as meaningful. In a meaningful loop, an increase in one variable will lead to a congruent change in the other variables, as represented by the direction of the associations. Examples of meaningful loops are a three-node or four-node loop with all positive associations, or a three-node loop with two positive and one negative associations.

## 3. Results

### 3.1. Correlations of children’s characteristics with BMI throughout childhood and genotype

Out of the 650 children with genotype data, 310 (47.7%) are females and 340 (52.3%) are males. The parent-reported race/ethnicity is Hipanic, Latino or Other race for 261 children (40.2%), Black for 250 (38.5%), and White for 139 (21.4%). 231 (35.5%) of the participants were enrolled at the San Diego study site, 169 (26%) at the Boston study site, and 250 (38.5%) at the St. Louis study site (Table 1). The mean BMI across the participants is 17 for age 2; 16.6 for age 3; 16.4 for age 4; 16.4 for age 5; 16.8 for age 6; 17.2 for age 7; and 18 for age 8. The median BMI percentile across the participants is 55.5 for age 2; 68.2 for age 3; 70.4 for age 4; 69.8 for age 5; 71.4 for age 6; 69.1 for age 7; and 72.1 for age 8. The BMI distribution is skewed towards the overweight category (**Figure *1***). To define possible confounders for our downstream analysis, we looked for associations between the children’s BMI and genotype first five principal components and their sex, race and ethnicity. We found significant associations (p-value < 0.05) between several time points (ages 5, 6, and 7) of BMI percentiles and the children’s sex. We found a weak association between the children’s BMI at age 2 and their sex. However, sex is not significantly associated with any of the children’s first five genotype principal components. Children’s race and ethnicity are significantly associated with their BMI at ages 2, and 4, and weakly associated at ages 5, and 8. On the other hand, race and ethnicity are significantly associated with the first four genotype principal components. The study site is associated with the children’s BMI at ages 1, 5, and 6, and weakly associated at age 8. Finally, the study site is significantly associated with the first four genotype principal components, and weakly associated with the fifth. Therefore, we adjusted our downstream analyses for race/ethnicity, sex, and study site.

**Table 1.** ***: p-value < 0.001; **: p-value < 0.01; *: p-value < 0.05; .: p-value < 0.1.

|  |  |  |  | BMI PERCENTILES |  |  |  |  |  |  |  | GENETIC PRINCIPAL COMPONENTS |  |  |  |
| --- | --- | --- | --- | --- | --- | --- | --- | --- | --- | --- | --- | --- | --- | --- | --- |
|  |  | N | % of cohort | Y2 | Y3 | Y4 | Y5 | Y6 | Y7 | Y8 | PC1 | PC2 | PC3 | PC4 | PC5 |
| SEX | Female | 310 | 47.7 |  |  |  |  |  |  |  |  |  |  |  |  |
|  | Male | 340 | 52.3 | . | ns | ns | * | * | * | ns | ns | ns | ns | ns | ns |
| RACE/<br>ETHNICITY | Hispanic/Latino or Other | 261 | 40.2 |  |  |  |  |  |  |  |  |  |  |  |  |
|  | Black | 250 | 38.5 | *** | ns | * | . | ns | ns | . | *** | *** | * | * | ns |
|  | White | 139 | 21.4 |  |  |  |  |  |  |  |  |  |  |  |  |
| STUDY SITE | San Diego | 231 | 35.5 |  |  |  |  |  |  |  |  |  |  |  |  |
|  | Boston | 169 | 26 | *** | ns | ns | * | * | ns | . | *** | *** | *** | ** | . |
|  | St. Louis | 250 | 38.5 |  |  |  |  |  |  |  |  |  |  |  |  |

**Figure 1.**
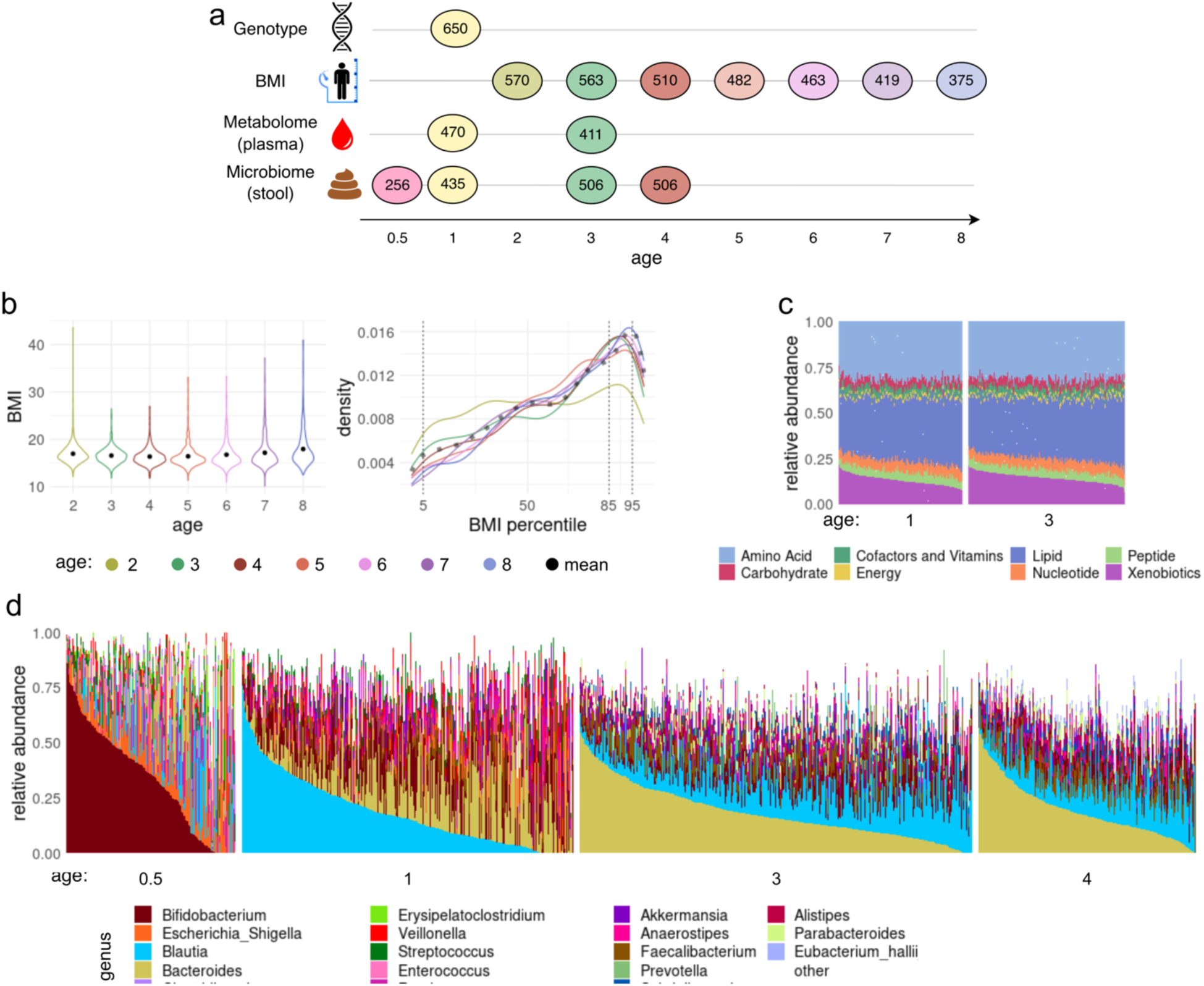
Children’s characteristics. **a.** Longitudinal data availability. Quantities in the panel represent the number of available samples of each different data type, at every timepoint between 0.5 to 8 years old. **b.** Distribution of child BMI per age (left), and of BMI percentiles per age (right). Black dots in the left panel represent the mean BMI per age, and black dotted line in the right panel represents the mean BMI percentile of all children at all ages available Vertical thin dotted lines on the right panel delimit the BMI categories regions: BMI under the 5th percentile is considered underweight; between the 5^th^ and 85^th^ percentiles is considered normal weight; between the 85^th^ and 95^th^ percentiles is considered over-weight; above the 95^th^ percentile is considered obesity. For all ages, the BMI is skewed towards the right and peaks at the overweight region. **c.** Metabolites abundance at ages 1 and 3. Colors represent the sum of the relative abundance of all metabolites in each of the 8 metabolite classes. Every vertical bar represents an available sample of plasma metabolites for the corresponding age. The metabolite composition is dominated by lipids and amino acids. Children’s metabolome tended to be relatively stable between the two timepoints with the difference that slightly more Xenobiotics were present at age 3 relative to age 1. **d.** Microbiome composition of children at different ages. The colors represent the 10 genera with highest mean relative abundance per age group across all children. Every vertical bar represents an available stool for the corresponding age. As children grow, their microbiomes tended to be richer, and the composition tended to be more even. The infant’s microbiome at age 0.5 tended to be dominated by the genus *Bifidobacterium*, and by *Blautia* at age 1. At age 3 and 4 the microbiome was dominated by *Bacteroides* and, was more similar than between any other two timepoints.

### 3.2. Association study of selected genes and gut microbiome identifies associations in the FHIT gene

To validate previously found genotype associations with the gut microbiome, we performed a targeted microbiome-genome association study on a pool of 12 genes and the alpha diversity and composition principal components of microbiome samples at ages 0.5, 1, 3, and 4 years. A Bonferroni-corrected significance level was established at p-value < 2.29 × 10^−6^, and a suggestive level at p-value < 10^−5^. We only found associations at the significant or suggestive level with variants in the FHIT gene (**Figure 2**.a). In total, four SNPs in the FHIT gene were associated with microbiome features: rs293602 is associated at the suggestive level with microbiome composition at age 0.5 PC1 (p-value < 4.5^-6^), and very close to the Bonferroni-corrected significance level; rs6781046 and rs1040338 are significantly associated with microbiome composition at age 3 PC2 (p-value < 2.2^-7^and p-value < 1.8^-6^, respectively; these two SNPs are located within 7430bp distance of each other and their minor allele count is highly correlated); and rs13086424 is significantly associated with microbiome richness at age 0.5 (p-value < 2.2^-6^). Because the association of rs293602 with the microbiome composition PC1 at age 0.5 is very close to the significance level, and above the suggestive level, we include it in the set of SNPs of interest. These four SNPs constitute the first part of our set of SNPs of interest. All the SNPs of interest identified at this point are located around the region 60Mbp (within the FHIT gene in chromosome 3) (**Figure 2**.c).

**Figure 2.**
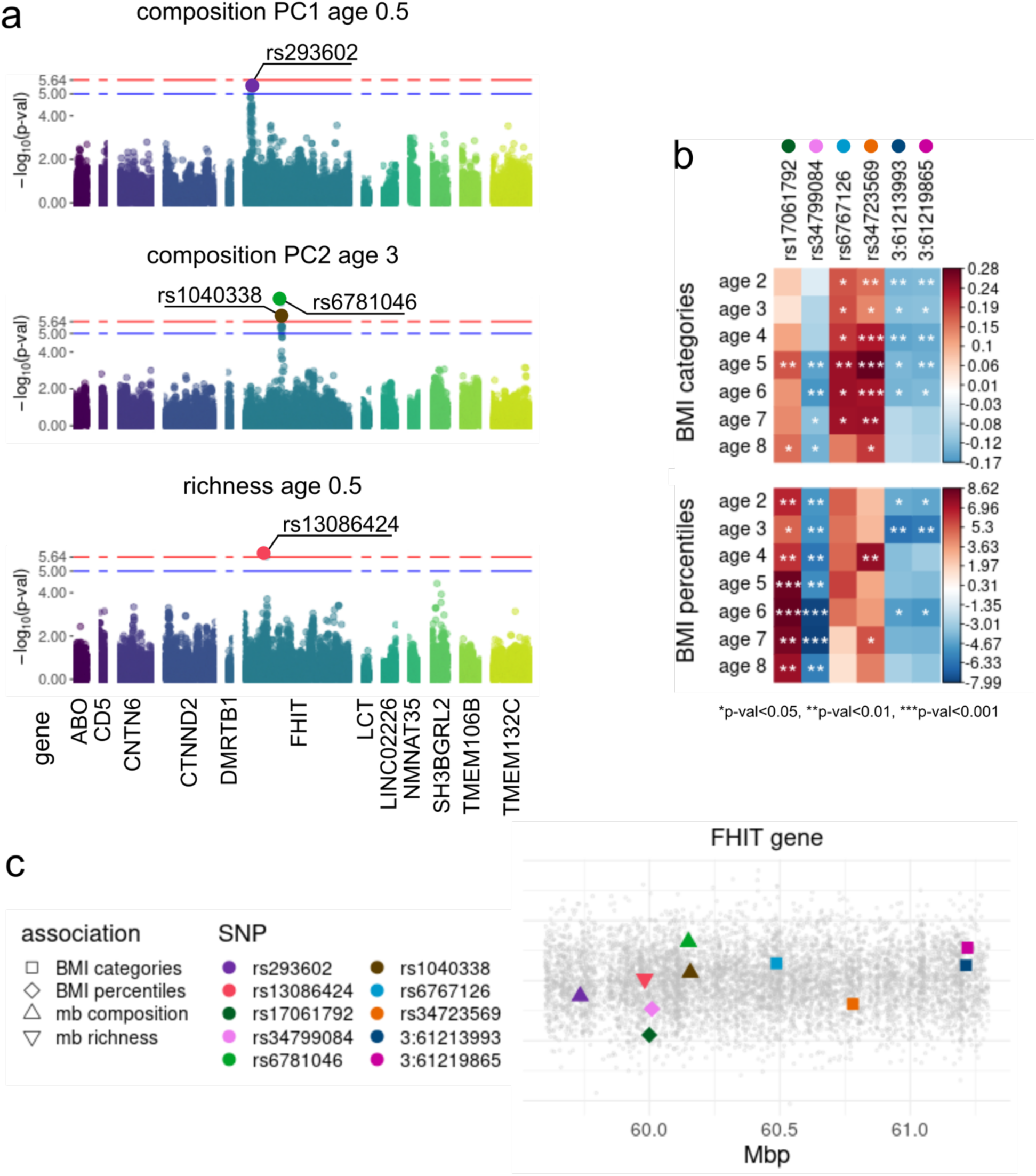
SNPs in the FHIT gene are associated with microbiome features and BMI. **a.** Four SNPs in the FHIT gene are associated with microbiome features. At the suggestive level(p-value < 10^−5^), rs293602 is associated with microbiome composition at age 0.5 PC1. At the adjusted GWAS significant level (p-value < 2.29 × 10^−6^), rs6781046 and rs1040338 are associated with microbiome composition at age 3 PC2, and rs13086424. **b.** Six additional SNPs in the FHIT gene are significantly (p<0.01) and consistently (for at least 5 consecutive timepoints) associated with BMI measurements (percentiles and categories). Heatmap shades represent covariate adjusted linear models coefficients. BMI categories are positively associated with the minor allele frequency in rs17061792, and negatively with that of rs34799084. BMI percentiles are positively associated with the minor allele frequency in rs6767126 and rs34723569, and negatively with that of 3:61213993 and 3:61219865. **c.** Locations of the SNPs associated with microbiome features and BMI measurements in the FHIT gene. Colors represent different SNPs, and shapes represent the associated variable. A small region around 60Mbp includes SNPs associated with microbiome features and a BMI measurement.

### 3.3. Loci in the FHIT gene are associated with childreńs BMI throughout childhood

To identify additional candidate loci for our downstream analysis, we looked for SNPs in the FHIT gene that are associated with child BMI measurements throughout childhood. Therefore, we ran covariate-adjusted linear regression on the minor allele count of the FHIT gene SNPs with respect to the BMI measurements at every age. We call an association consistent when it is statistically significant (p-value < 0.05) for at least 5 consecutive time points of one BMI measurement (BMI categories or percentiles), and the association maintains the same direction for all time points across both measurements. In total, 5 consistent associations were found (**Figure 2**.b): The frequency of allele T in rs17061792, and of allele TAA in rs34799084 are positively and negatively associated with the children’s BMI percentiles, respectively (all p-values for ages 2 through 8 < 0.05 and < 0.01 respectively); the frequency of allele C in rs6767126 and of allele A in rs34723569 are positively associated with the children’s BMI categories (p-values < 0.05 for ages 2 through 7, and 2 through 8 respectively); and the frequency of allele T and G in 3:61213993 and 3:61219865, respectively, is negatively associated with the BMI percentiles (p-values < 0.05 for ages 2 through 6). **Figure 2**.c shows the locations of the FHIT gene SNPs consistently associated with child BMI which, along with SNPs associated with microbiome features, complete our set of SNPs of interest. Notably, the two SNPs consistently associated with the BMI percentiles are in very close proximity of those associated with microbiome composition and richness within the FHIT gene.

### 3.4. The abundances of *Blautia*, *Ruminoccocus gnavus* and other genera are associated with variants in the FHIT gene

Next, we investigated how genetic variants in the FHIT gene SNPs of interest and child BMI relate to the relative abundance of individual taxa in their gut microbiome. To do so, we first looked for differentially abundant taxa with respect to the minor allele counts (MAC) of the SNPs of interest and with respect to the BMI measurements at every age through independent MaAsLin analyses at the microbiome genus level. Then, we fitted mixed models of the relationship between the genotype and BMI measurements, to keep track of the repeated measurements per child.

We found that the relative abundance of six genera at three different time points are significantly associated (MaAsLin q-value < 0.05) with the minor allele count of 6 of the 10 SNPs of interest (**Figure 3.**a). Most of the associations are with taxa at age 3 years: we found positive associations (a specific genus is enriched in children with 2 minor alleles at a specific locus, and depleted in children with zero minor alleles at that same locus) between the abundance of *Blautia* and the minor allele counts of SNPs rs6781046 and rs1040338 (both previously associated with microbiome composition), and between the abundance of *Collinsella* and SNP rs6767126 (previously associated with BMI categories). The association between *Blautia* and rs1040338 is the strongest among all the differential abundance results; the minor allele counts of SNPs rs6781046 and rs1040338 are highly correlated (SI Figure). At age 3 years, we also found a negative association between the abundance of *Allistipes* and *Agathobacter*, and the minor allele counts of SNPs 3:61213993 and 3:61219865 (both previously associated with BMI categories); the minor allele counts of these two SNPs are also highly correlated (SI Figure). At age 4 years, we found that *Ruminoccocus gnavus* group is positively associated with the minor allele count of 3:61213993 and 3:61219865; these are the second and third strongest associations that we found. The least strong association is between the abundance of *UBA1819* at age 0.5 years and the minor allele count of SNP rs13086424 (previously associated with microbiome richness). **Figure 3.**b shows the relative abundance of the differentially abundant genera with respect to the minor allele count of the associated SNPs. All genera, except for *Blautia*, were absent in a large proportion of children at the significantly associated ages (87.4% of samples with zero values for *UBA1819*; 0.6% for *Blauti*a; 46.2% for *Collinsella*; 22.5% for *Allistipes*; 34.7% for *Agathobacter*; and 64.3% for *Ruminoccocus gnavus* group), but their relative abundance is substantial across the samples (0.002 mean relative abundance for *UBA1819*; 0.15 for *Blauti*a; 0.012 for *Collinsella*; 0.021 for *Allistipes*; 0.014 for *Agathobacter*; and 0.008 for *Ruminoccocus gnavus* group) except for *Collinsella* at age 3 (0.075 maximum relative abundance) and *UBA1819* at age 0.5 (0.068 maximum relative abundance). We conclude that their associations with the corresponding SNPs are less robust than the others we observed, though statistically significant. The six genera identified in this step (*UBA1819*, *Blautia*, *Collinsella*, *Allistipes*, *Agathobacter*, and *Ruminoccocus gnavus* group) are referred to in the following as the genera of interest.

**Figure 3.**
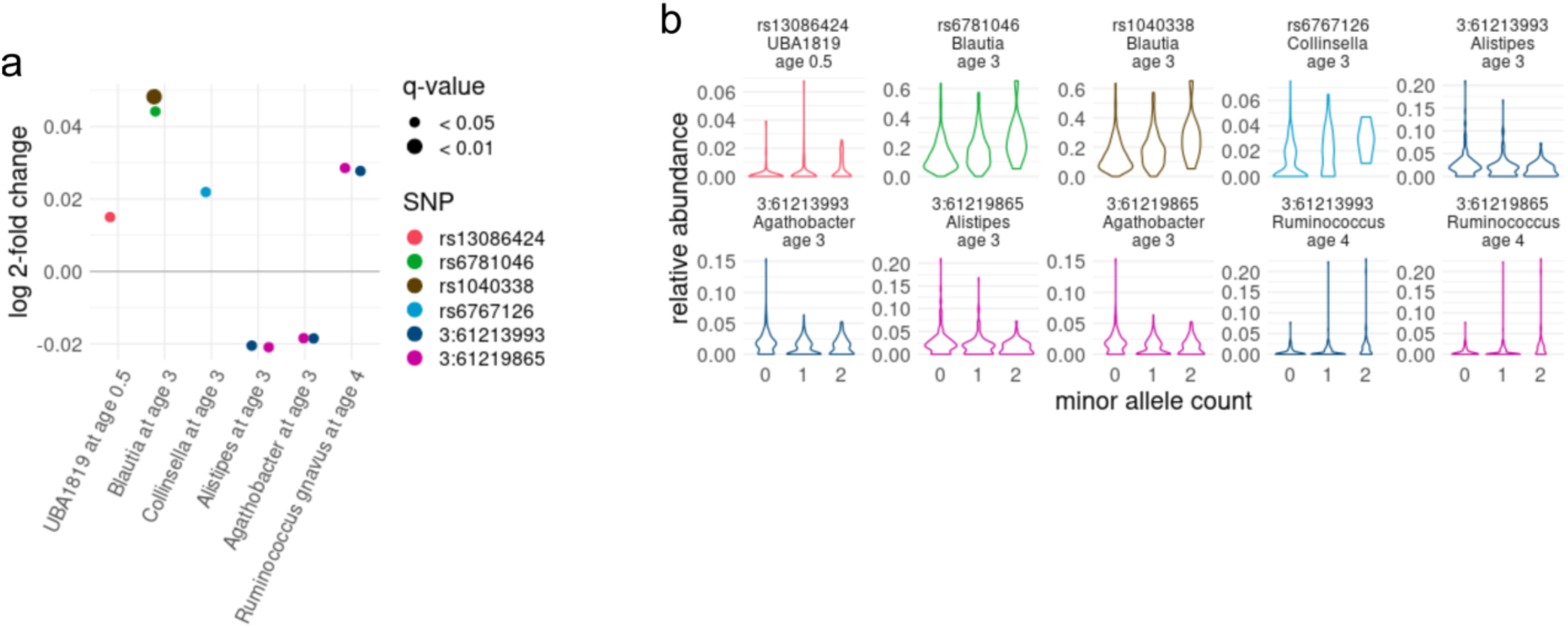
Six microbiome genera at ages 0.5, 3, and 4 are differentially abundant with respect to the minor allele count (MAC) of the selected SNPs (Maaslin2 q-val < 0.05, MAC (0,1,2) is treated as a continuous variable). **a.** The strongest associations is between rs6781046 and rs1040338 and *Blautia* at age 3, the first having the also the strongest statistical significance, followed by 3:61213993 and 3:61219865 and *Ruminococcus gnavus* group at age 4. These associations have a positive coefficient. The rest of the associations are between *UBA1819* at age 0.5 and rs13086424, *Collinsella* at age 3 and rs6767126, and *Allistipes* and *Agathobacter* at age 3. The first two have positive coefficients while the latter two have negative coefficients. **b.** The relationship between the relative abundance of the differentially abundant bacteria and the minor allele count of the corresponding SNPs follows the coefficient sign pattern showed in panel a. Colors in this panel correspond to the SNPs in panel a. Highly correlated pairs of SNPs (rs6781046 and rs1040338, and rs6781046 and rs1040338) show almost identical patterns. All genera, except for *Blautia*, have an important number of zero values. However, only *UBA1819* at age 0.5, and *Collinsella* at age 3 have overall low relative abundance. Out of the 6 genera associated with genotype, *UBA1819* presents the least robust association.

Surprisingly, despite our prior finding of significant associations between BMI measurements and the minor allele count of SNPs linked to differential abundance in certain genera, we did not observe any associations between gut taxa at the genus level and BMI measurements (SI Figure).

### 3.5. Metabolic pathways are associated with BMI measurements

To investigate the interconnections between the children’s genotype, microbiome and their metabolome in the context of BMI measurements, we looked for associations between the relative abundance of metabolites and child BMI measurements. We found a total of 25 plasma metabolites (9 amino acids, 2 carbohydrates, 9 lipids, 2 nucleotides and 3 xenobiotics) whose relative abundance is significantly associated (p-values < 0.01) with either BMI percentile, BMI category, or both (**Figure 4**.a). These 25 metabolites span 21 metabolic pathways that we carried forward in downstream analyses, in which we will refer to these pathways as pathways of interest. Two forms of Sphingolipid Sphingomyelin, and CMPF (a fatty acid in the dicarboxylate metabolism), are associated with both BMI measurements with similar significance. Most of the other associations are between plasma metabolite relative abundances and BMI percentiles; the two exceptions with the strongest associations are another form of sphingomyelin and the xenobiotic benzoate 4-ethylphenyl sulfate, both of which are positively associated with BMI categories. The Xenobiotic 4-ethylphenyl sulfate, along with the amino acid Cysteine s-sulfate in the Methione, Cysteine, SAM and Taurine metabolism pathway (Sulfur Amino Acids) and the three forms of Sphingomyelin are the only metabolites to have positive associations with BMI measurements; all the other observed associations are negative. While we observed few associations between carbohydrates and BMI measurements, arabytol/xylitol has the strongest overall association that we found. This carbohydrate is in the pentose pathway and is negatively associated with BMI percentiles. Sphingolipid metabolism is the pathway with the higher number of associations (n=3).

**Figure 4.**
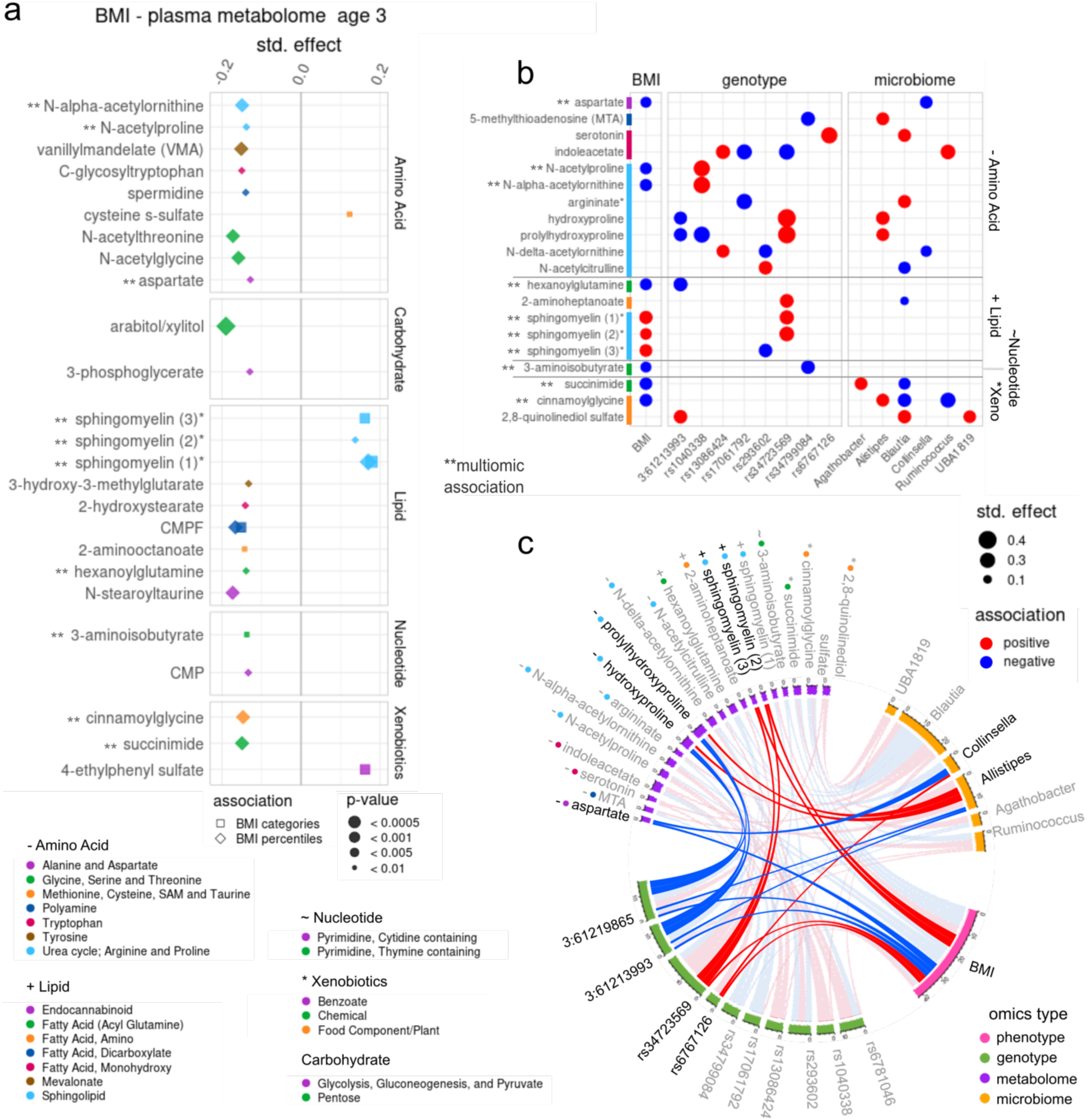
Pathways in the plasma metabolome are simultaneously associated with BMI measurements and selected microbiome and genotype features. Linear models are used to establish associations between the metabolites’ abundance and the different omics variables (BMI percentiles or categories, genotype, or microbiome features). The coefficient is standardized as β*sd(explanatory variable measurement in all children)/sd(metabolite abundance in all children) to allow magnitude comparison (std. effect in the panels). **a.** Metabolome associations with BMI measurements. Associations with BMI percentiles are more prevalent than with BMI categories, but two lipids have significant associations with both. Amino acids and Lipids are the metabolite classes with most significant associations (n = 9, respectively). Cysteine s-sulfate in the Methione, Cysteine, SAM and Taurine metabolism pathway (Sulfur Amino Acids) is the only amino acid to have a positive association with BMI (categories), while all the other found associations are negative. The overall strongest association (negative) is between the carbohydrate arabitol/xylitol in the pentose pathway, and BMI percentiles, while the most consistent pathway association is between three lipids in the Sphingolipid pathway and both BMI measurements. In total, we identified 21 metabolic pathways, in 5 metabolite classes associated with BMI measurements. **b.** Metabolites simultaneously associated with two other data types (BMI measurements, genotype, or microbiome features). In total, we found 20 metabolites in 10 pathways and 4 metabolite classes that are associated with features in two other types of data. The most recurrent are associations between metabolites, genotype, and microbiome (n=9) and, in particular, with metabolites in the Urea cycle and Arginine and proline metabolism (n=4). Many of the metabolite associations with genotype features involve more than one SNP per metabolite. This is also observed in the metabolite microbiome associations where at least 2 genera are associated with each metabolite. **c.** Network representation of all the associations found. Nodes are genetic, metabolome, microbiome or BMI variables, and links represent significant associations (red for positive and blue for negative). This undirected network representation allows to analyze the interconnections simultaneously. We found two three-node and one four-node interconnections with coherent directional association i.e., an increase in one variable corresponds adequately to changes in others: 1) SNPs *3:61213993 and 3:61219865,* amino acids hydroxyproline and prolyhydroxyproline, and genus *Allistipes;* 2) SNP *rs34723559*, lipid *sphingomyelin,* and BMI; and 3) SNP rs6767126, genus *Collinsella*, BMI, and aspartate aminotransferase.

Using the 21 metabolic pathways of interest, we searched for associations between the relative abundance of all their metabolites and both the minor allele count of the SNPs of interest, and the relative abundance of genera of interest. Focusing on the metabolites that have a simultaneous association with at least one variable of interest in each of two different types of data (BMI and genotype, genotype and microbiome, or microbiome and BMI), we find 20 matches with statistical significance (p-value < 0.01) among Amino acids (n=11), Lipids (n=5), Nucleotides (n=1), and Xenobiotics (n=3) (**Figure 4**.b). These 20 metabolites span 10 of the 21 metabolic pathways of interest. Most of them are associationed with genotype and microbiome (n=10), followed by BMI and genotype (n=7) and, lastly, with BMI and microbiome (n=3).

### 3.6. Integrated network analysis

Finally to analyze all the results that we obtained in the previous steps in an integrated manner, we built the network in **Figure 4**.c, where every node represents one metabolite, SNP, genera, or BMI measurement, and the links represent associations between them. We define a loop as a closed connection between three or four nodes of different omics types. We deem a loop as meaningful if the direction of the associations is coherent (an increase in one variable corresponds adequately to changes in others). For example, a three-node loop is meaningful if the three nodes are either connected by all positive links, or two negative and one positive link. We found 9 three-node loops, and 7 four-node loops of which 3 and 1 were meaningful, respectively (highlighted in in **Figure 4**.c). The first three-node loop has all positive links and is formed by the association between BMI, SNP rs34723569 and two forms of sphingomyelin. The other two three node loops include the highly correlated SNPs 3:61219865 and 3:61213993, which have a negative association with prolyhydroxyproline and hydroxyproline respectively. These two amino acids are positively associated with the relative abundance of *Allistipes*, which is negatively associated with both 3:61213993 and 3:61219865. Importantly, 3:61213993 and 3:61219865 have a positive association with BMI. The meaningful four node loop is formed by the positive associations of the MAC in rs6767126 with both the relative abundance of *Collinsella* and the BMI, and the negative associations of the relative abundance of the amino acid aspartate and both *Collinsella* and BMI.

## 4. Discussion

Whether and to what extent genotype contributes to shaping the human microbiome is a long withstanding open question in the field. One major challenge in addressing it is the large multiple testing burden associated with GWAS and the large cohorts required to draw significant conclusions. Similarly, the integration of multiple types of omics data presents the challenge of high dimensionality and the problem of selecting variables from a very large pool. We address these problems by reviewing existing results in the microbiome-GWAS literature and taking a knowledge-based targeted sequential approach to the variable selection process.

By selecting a pool of 12 genes that are known to be associated with microbiome features [21] for our microbiome-GWAS, we aim to test the validity of existing results in the developing infant and early childhood microbiome rather than find new associations. We can therefore establish a Bonferroni-corrected significance level of 2.29 × 10^−6^, which allows for less stringent testing than classical GWAS. The alpha diversity measurements and the dimensionality reduction that robust Aitchison PCA [50] provides for the microbiome composition allowed us to narrow down our downstream analysis to 10 SNPs in the FHIT gene that are associated with either microbiome diversity, composition, or BMI. Variants in the FHIT gene have been iteratively found to be associated with obesity in adults and children [53–55] and with components of the fecal microbiome associated with visceral fat [56]. This knowledge, paired with our results, suggest that the identified variants might be implicated in determining BMI. Concordantly, the genus *Blautia*, which is known to be negatively associated with visceral fat and overweight/obesity in adults and children [57–60], is the most heavily loaded on the principal component with the strongest association (PC2 at age 3). The downstream analysis included three more steps: 1) identification of 6 genera (*UBA1819*, *Blautia*, *Collinsella*, *Allistipes*, *Agathobacter* and *Ruminoccocus gnavus*) that are differentially abundant with respect to the children’s genotype in the identified SNPs; 2) identification of metabolic pathways that are significantly associated with child BMI; and 3) search for simultaneous associations of the relative abundance of individual metabolites in the identified pathways and the relative abundance of identified genera and the children’s genotype. The main result of our analyses is the discovery of three closed-loop multi-omic associations: two interconnections between three variables each of a different omics type, and one between four variables of different omics types. These interconnections may shed some light on the role that a genotype-microbiome-metabolome axis plays in conditions such as obesity and type 2 diabetes.

First, we found an association between a higher minor allele count in the FHIT SNP rs34723559, the enrichment of two forms of sphingomyelin, and increased BMI; higher BMI is also associated with a higher minor allele count in rs34723559. Because sphingomyelin has been found to inhibit insulin action in humans [61], and in mice a reduction in plasma sphingomyelin led to resistance to obesity when fed a high fat diet and to higher sensitivity to insulin [62], this interconnection suggests that having a homozygous minor genotype at the rs34723559 locus might represent a higher risk of obesity and obesity related complications such as insulin resistance or type 2 diabetes.

Second, we found a triple interconnection including a low minor allele count in the highly correlated FHIT gene SNPs 3:61213993 and 3:61219865 associated with the enrichment of two amino acids in the urea cycle: hydroxyproline and prolyhydroxyproline, and of the genus *Allistipes*; hydroxyproline, prolyhydroxyproline, and *Allistipes* are positively associated. Hydroxyproline is known to inhibit the degradation of incretin hormones by dipeptidyl peptidase (DPPIV) [63,64] and, therefore, promote the maintenance of blood sugar at normal levels. Indeed, DPPIV inhibitors have been proven to be effective treatment against Type 2 diabetes without inducing weight gain or loss effects as other available treatments [65,66]. On the other hand, the strain *Allistipes putredinis* has been found depleted in patients with obesity [8], but other *Allistipes* strains have been found to have either protective or pathogenic effects in several cardiovascular and inflammation related conditions [67]. Interestingly, we found that a low minor allele count in 3:61213993 and 3:61219865 is also associated with increased child BMI. More in-depth studies are needed to establish more precise conclusions with respect to this interconnection, especially regarding the specific *Allistipes* strains involved in the associations. Metagenomic sequencing of stool samples could provide such detailed strain information.

Third, the only interconnection that includes all four omics data types includes a homozygous minor genotype in the FHIT SNP rs6767126, which is associated with enrichment of *Collinsella* and increased child BMI. *Collinsella* is considered a pro-inflammatory genus and, consistent with our result, there is evidence of its relative abundance decreasing during a weight loss program [68]. Moreover, an enrichment of *Collinsella* has been found to be associated with type 2 diabetes [69] and compromised liver function [70]. In our cohort, a higher child BMI and enriched *Collinsella* are simultaneously associated with a lower relative abundance of the amino acid aspartate. Interestingly, there is evidence of the heritability of aspartate aminotransferase [71], and it is classically considered a biomarker of liver disease [72], though it is a higher abundance that is considered indicative of decreased liver function. However, in a study done in obese women, the significance of the BMI and aspartate relative abundance disappeared after correcting for dietary intake [73]. Considering that many genome-wide associations of serum aspartate modified by BMI have been reported, but none on the FHIT gene [74], more investigations are necessary to determine the role of aminotransferase metabolites in the context of child obesity and diabetes risk.

In summary, we have presented evidence that the genotype influences the gut microbiome and blood metabolome composition, with relevance to phenotypic characteristics such as BMI. Notably, we have validated the known association of genetic variants with microbial genera previously found in adults, in children whose microbiomes are in a developmental stage. Additionally, we have discussed the potential implications of these associations on negative health outcomes related to child obesity and its complications such as insulin resistance and type 2 diabetes. The variables involved in the four closed loops that we found can be understood as new potential markers for tracking the risk of these conditions. Moreover, the interconnections between them can be interpreted as redundancies that offer the possibility of more robust health screening that could help design early interventions and ensure healthy development of children.

### Ethical Statement and Institutional Board Review

The VDAART study protocol was approved by the institutional review boards at each participating institution and at Brigham and Women’s Hospital. The study was conducted in accordance with the Declaration of Helsinki.

### Informed Consent Statement

Informed consent was obtained from all subjects involved in the study.

## Data Availability

Genotype, Microbiome sequencing, metabolome and anthropometric measurements data are part of the ECHO consortium and ECHO consortium members can obtain the data directly from the ECHO DCC or, for those not part of ECHO, directly from the authors.

## Funding

VDAART was funded by U01HL091528 from the National Heart, Lung, and Blood Institute. Additional funding came from NIH grants K08HL148178, R01HL108818, T32HL742742, and ECHO grant OD023268.

## Notes

### Competing Interest Statement

The authors have declared no competing interest.

### Clinical Trial

NCT00920621

### Author Declarations

Ethics commitee of Brigham and Women's Hospital gave ethical approval for this work.

